# Protocol for the Redefining Maternal Anemia in Pregnancy and Postpartum (ReMAPP) study: A multisite, international, population-based cohort study to establish global hemoglobin thresholds for maternal anemia

**DOI:** 10.1101/2024.11.06.24316823

**Authors:** Zahra Hoodbhoy, Aneeta Hotwani, Fyezah Jehan, Amna Khan, Imran Nisar, Nida Yazdani, Santosh Joseph Benjamin, Anne George Cherian, Venkata Raghava Mohan, Sunitha Varghese, Balakrishnan Vijayalekshmi, Blair J. Wylie, Leena Chatterjee, Arjun Dang, R Venketeshwar, Sasha G. Baumann, Christopher Mores, Qing Pan, Emily R. Smith, Christopher R. Sudfeld, Victor Akelo, □Winnie K. Mwebia, Kephas Otieno, Gregory Ouma, Harun Owuor, Joyce Were, Dennis Adu-Gyasi, Veronica Agyemang, Kwaku Poku Asante, Sam Newton, Charlotte Tawiah, Arun Singh Jadaun, Sarmila Mazumder, Neeraj Sharma, Lynda G. Ugwu, Amma Benneh-Akwasi Kuma, Bethany Freeman, Margaret P. Kasaro, Felistas M. Mbewe, Humphrey Mwape, Rachel S. Resop, M. Bridget Spelke

## Abstract

**Background:** Anemia affects one in three pregnant women worldwide, with the greatest burden in South Asia and sub-Saharan Africa. During pregnancy, anemia has been linked to an increased risk of adverse maternal and neonatal health outcomes. Despite widespread recognition that anemia can complicate pregnancy, critical gaps persist in our understanding of the specific causes of maternal anemia and the cutoffs used to diagnose anemia in each trimester and in the postpartum period.

**Methods and analysis:** The Redefining Maternal Anemia in Pregnancy and Postpartum (ReMAPP) study is a multisite, prospective, cohort study nested within the Pregnancy Risk, Infant Surveillance, and Measurement Alliance (PRISMA) Maternal and Newborn Health study. Research sites are located in Kenya, Ghana, Zambia, India, and Pakistan. Participants are up to 12,000 pregnant women who provide serial venous blood samples for hemoglobin assessment at five time points: at <20 weeks, 20 weeks, 28 weeks, and 36 weeks gestation and at six weeks postpartum. We will use two analytical approaches to estimate hemoglobin thresholds for defining anemia: (1) clinical decision limits for cutoffs in each trimester and at six weeks postpartum based on associations of hemoglobin levels with adverse maternal, fetal, and neonatal health outcomes and (2) reference limits for gestational-week-specific cutoffs and at six weeks postpartum for mild, moderate, and severe anemia based on tail statistical percentiles of hemoglobin values in a reference (i.e., clinically healthy) subpopulation. We will also conduct biomarker-intensive testing among a sub-sample of participants in each trimester to explore underlying contributing factors of maternal anemia.

**Ethics and dissemination:** The study received local and national ethical approvals from all participating institutions. Findings from multisite analyses will be published among open-access, peer-reviewed journals and disseminated with local, national, and international partners.

**Registration:** ClinicalTrials.gov (ID: PRISMA-MNH 2022; NCT05904145)

**Strengths and limitations:**

- Novel study design to allow multiple analytical approaches (clinical decision limits and reference limits) in the same population to establish hemoglobin thresholds.
- Use of gold standard methods and external quality assurance programs to ensure harmonized hemoglobin measurement across sites.
- Inclusion of biomarker-intensive study arm to examine the etiology of anemia among pregnant women.
- All data is contributed by populations historically underrepresented in research in low- and middle-income countries.

## INTRODUCTION

Anemia is a common health condition characterized by low blood hemoglobin concentration and/or low red blood cell count. Women, particularly those who are pregnant and lactating, are physiologically at a higher risk than men of being anemic [1,2]. An estimated 36% of pregnant women globally are anemic, with the highest prevalence in South Asia (48%) and West and Central Africa (52%) [3]. Although there are many causes of anemia, iron deficiency is estimated to account for roughly half of anemia cases in women [4]. Other contributing causes of anemia include infectious diseases (e.g., hookworm, malaria, schistosomiasis, HIV), gynecologic conditions (e.g., abnormal uterine bleeding, hemorrhage, uterine fibroids), other micronutrient deficiencies (e.g. folate, vitamin B12), thyroid dysfunction, chronic kidney disease, hemoglobinopathies like sickle cell disease or thalassemia, and environmental exposures such as lead or arsenic [1,2,5–8]. Importantly, causes of anemia vary widely by geography, life stage, and age.

During pregnancy, abnormal hemoglobin levels are linked to an increased risk of adverse maternal, fetal and neonatal health outcomes. Specifically, maternal anemia (i.e. low hemoglobin) can increase risk of postpartum hemorrhage, preeclampsia, maternal mortality, postpartum maternal depression, preterm delivery, low birth weight, small for gestational age, stillbirth, neonatal mortality, and poor infant brain structural development [9–13]. The likelihood of these outcomes depends on the case severity, gestational timing onset and duration of anemia [11,12]. Multiple studies have also observed an association between high hemoglobin concentration and pregnancy complications, though there are no standard thresholds to define excess hemoglobin [11,14–16]. In a systematic review and meta-analysis by Young et al. (2023), hemoglobin levels greater than 13.0 g/dL during pregnancy were associated with increased risk of small-for-gestational-age, stillbirth, (very) low birth weight, preterm birth, gestational diabetes, preeclampsia, and maternal mortality [17].

A growing body of evidence over the last decade suggests that the hemoglobin cutoffs used to diagnose anemia during pregnancy and the postpartum period should be reevaluated in consideration of data from low- and middle-income countries (LMICs) [11,18–20]. In 2017, a World Health Organization (WHO) technical consultation concluded that the current hemoglobin thresholds used in pregnancy were not supported by sufficient data, not proven to be linked to health outcomes, not representative of diverse geographies and ethnicities, and not necessarily applicable in the context of common gene mutations that affect hemoglobin function [21]. Although WHO released revised anemia definitions in 2024, the use of trimester-specific cutoffs during pregnancy and of non-pregnant adult cutoffs in the immediate postpartum period to diagnose anemia are not well-evidenced [22]. Similarly—given that causes of anemia are varied in severity, multifactorial, and regionally distinct—further research is needed to understand how anemia etiology differs for pregnant women, given that most research to date has focused on non-pregnant women [23].

Correct anemia identification carries significance not only for its prevention, diagnosis, and treatment at the individual level, but also for tracking progress toward global anemia reduction targets. Hemoglobin thresholds to diagnose anemia in pregnancy at all gestational ages were first established by the World Health Organization (WHO) in 1959 as <10.0 g/dL, then adapted in 1968 to <11.0 g/dL [24,25]. These definitions of anemia were based on a ‘normal range’ of hemoglobin values; wherein below the 2.5th percentile (−2 standard deviations (SDs) from the mean) is considered low hemoglobin (i.e. anemia) and above the 97.5th percentile (+2 SDs from the mean) is high hemoglobin. Data informing the ‘normal range’ was largely derived from four published studies in high-income countries (Norway, England, Wales, United States) with small sample sizes, not all of which included women [26–29]. In 1989, the Centers for Disease Control (CDC) set thresholds based on studies in Europe and the United States for each trimester: <11.0 g/dL in the first and third trimesters and <10.5 g/dL in the second trimester [30]. The WHO introduced thresholds for anemia severity in pregnant women in 2011, adopted the CDC trimester-specific thresholds in 2016, and added trimester-specific severity cutoffs in 2024 (**Table 1**) [22,31,32]. Notably, the WHO guidelines released in 2024 excluded data from LMICs in the pooled analysis that informed the update, citing that the unknown effect of infection and inflammation on hemoglobin coupled with their high prevalence in these settings precluded their inclusion [22].

**Table 1.** WHO (2024) thresholds for anemia for non-pregnant and pregnant women.

| Population | Hemoglobin threshold (g/dL) |  |  |  |
| --- | --- | --- | --- | --- |
|  | Any anemia | Mild anemia | Moderate anemia | Severe anemia |
| Non-pregnant women | <12.0 | 11.0 - 11.9 | 8.0 - 10.9 | <8.0 |
| Pregnant women | - | - | - | - |
| 1st trimester (≤13 weeks) | <11.0 | 10.0 - 10.9 | 7.0 - 9.9 | <7.0 |
| 2nd trimester (14-27 weeks) | <10.5 | 9.5 - 10.4 | 7.0 - 9.4 | <7.0 |
| 3rd trimester (≥28 weeks) | <11.0 | 10.0 - 10.9 | 7.0 - 9.9 | <7.0 |

We designed the Redefining Maternal Anemia in Pregnancy and Postpartum (ReMAPP) study to establish hemoglobin cutoffs using multiple analytical approaches for the diagnosis of anemia during pregnancy and within 42 days postpartum in LMIC populations. The specific aims are threefold: (1) to determine trimester-specific and postpartum hemoglobin thresholds for anemia diagnosis based on statistically significant increased risks of adverse maternal, fetal, and newborn health outcomes (i.e. decision limits); (2) to estimate gestational-week-specific and six-week postpartum hemoglobin thresholds for mild, moderate, and severe anemia diagnosis using the 5th and 2.5th statistical percentiles among a clinically healthy subpopulation (i.e. reference limits); (3) to describe the underlying contributing factors of anemia (i.e. etiology) in pregnancy in Ghana, Kenya, Zambia, Pakistan, and India.

## MATERIALS AND METHODS

### Study design

ReMAPP is a prospective cohort study in five countries. It is embedded within the Pregnancy Risk, Infant Surveillance, and Measurement Alliance (PRISMA) Maternal and Newborn Health study: a longitudinal, open cohort study that seeks to evaluate pregnancy risk and health and development for women and infants [33]. The population for ReMAPP is nested in the PRISMA study cohort. Enrollment for ReMAPP was initiated on a rolling basis by site and began between September 2022 and December 2023 (22 September 2022 in Pakistan, 15 December 2022 in Zambia, 28 December 2022 in Ghana, 14 April 2023 in Kenya, 20 June 2023 in India (Vellore), and 12 December 2023 in India (Hodal). Enrollment is expected to be completed by June 2025. As illustrated in **Figure 1**, further selection will be done from the primary cohort of enrolled participants to identify a clinically ‘healthy’ analytical subcohort and cross-sectional sub-samples for etiology assessment. Data from the primary cohort, analytical subcohort, and cross-sectional sample will be used to achieve aims one, two, and three, respectively.

**Figure 1.**
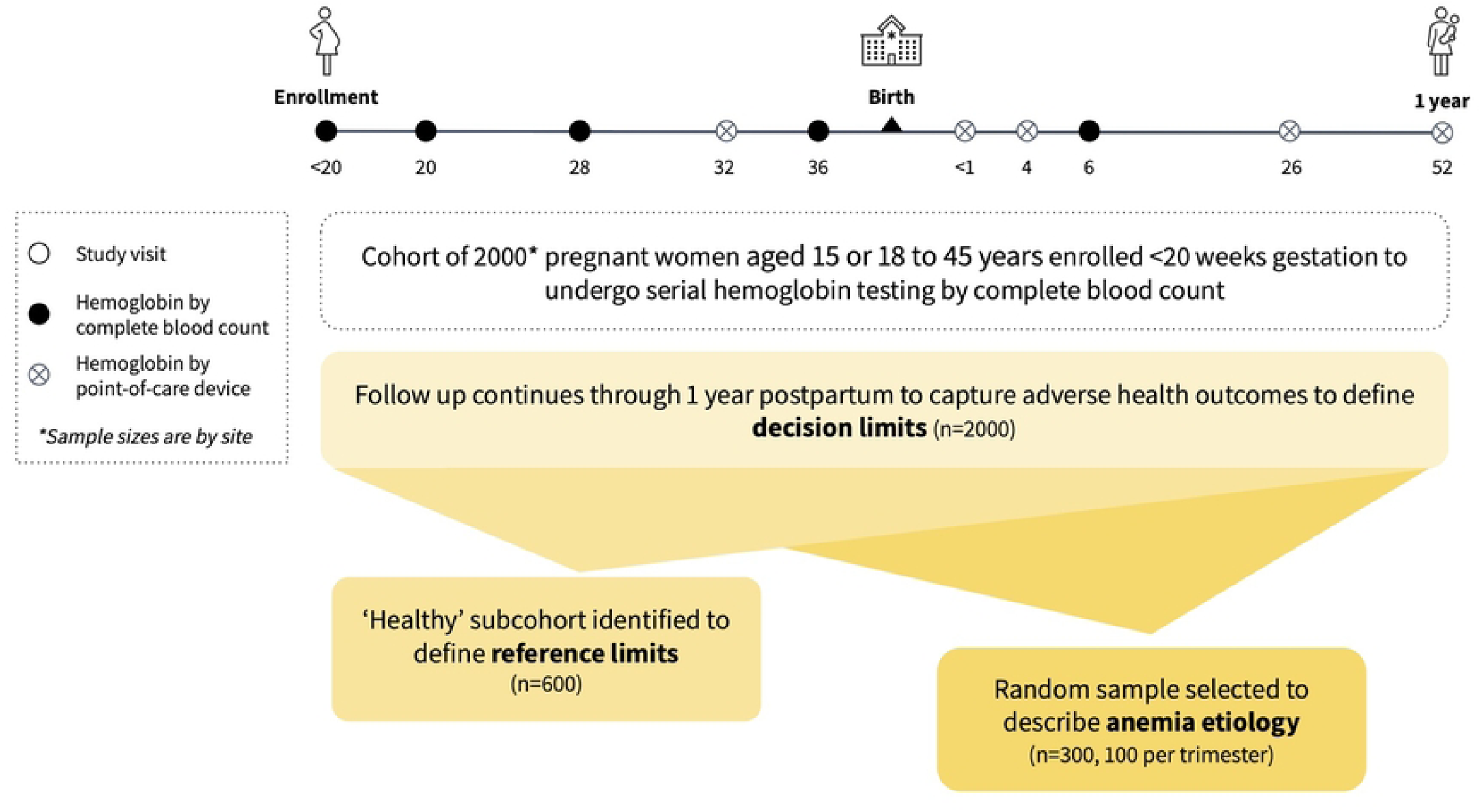
Nested ReMAPP study design with estimated 12,000 pregnancies in study cohort (n=1,650 to 2,000 each site)

### Study setting

This study focuses on countries in South Asia and sub-Saharan Africa with high anemia prevalence [34]. There are six research sites: Kintampo, Ghana; Kisumu and Siaya, Kenya; Lusaka, Zambia; Karachi, Pakistan; Vellore, India; and Hodal, India. Each site has identified and mapped geographic catchment areas, which have at least 1500 deliveries per year.

### Eligibility criteria and recruitment

Participants eligible for the ReMAPP study are pregnant women who live within the catchment area, are <45 years and meet country-specific minimum age requirements (Ghana: 15 years of age; Kenya: 18 years of age or emancipated minors; Pakistan: 15 years of age or emancipated minors; Zambia: 15 years of age; India: 18 years of age), have a viable intrauterine pregnancy, are less than 20 weeks gestation as verified via ultrasound, intend to stay in the study area until six weeks post-delivery, and who provide informed consent.

### Participant selection and sampling

All participants enrolled in the PRISMA study will be invited to participate in the ReMAPP study if they meet eligibility criteria. Enrollment will continue until the target sample size (n=1650 to 2000 per site) is achieved. These participants will form the ReMAPP primary cohort, and will undergo serial hemoglobin testing by complete blood count and be followed up at regular study visits through one year postpartum.

Upon enrollment, all ReMAPP participants will undergo screening based on predetermined criteria to obtain sociodemographic information, obstetric history, vital signs, anthropometric measurements, and laboratory results. If a participant meets the 21 screening criteria (**Box 1**), they will be included in the clinically ‘healthy’ analytical subcohort. Selected participants may also be excluded from the final ‘healthy’ analytical subcohort if any of the following occur during follow-up: multiple pregnancies previously not identified, severe conditions not initially evident including cancer, HIV, tuberculosis, or malaria, or severe pregnancy-related conditions requiring hospital admission including severe preeclampsia/eclampsia.

#### Box 1. Screening criteria for clinically healthy subcohort

1. Aged 18 to 34 years
2. Gestational age at enrollment <14 weeks
3. Pre-pregnancy or early pregnancy body mass index (BMI) of >18.5 and <30 kg/m^2^
4. Mid-upper arm circumference >23cm
5. Height ≥150 cm
6. Singleton pregnancy
7. Systolic blood pressure <140 mmHg and diastolic blood pressure <90 mmHg
8. No iron deficiency (serum ferritin >15 mcg/L adjusted for inflammation [35])
9. No subclinical inflammation (CRP≤5 mg/L and/or AGP≤1 g/L)
10. No hemoglobinopathies (SS, SC, SE, EE, CC, SD-Punjab, Sβthal, Eβthal, Cβthal, CD-Punjab, ED-Punjab, D-D-Punjab, D-Punjabβthal, Thalassemia major, Thalassemia intermedia, or Alpha thalassemia)
11. Normal glucose-6-phosphate dehydrogenase (≥6.1 U/g Hb)
12. No reported previous low birth weight delivery
13. No reported previous stillbirth
14. No reported previous unplanned cesarean delivery
15. No reported cigarette smoking, tobacco chewing, or betel nut use
16. No reported alcohol consumption during pregnancy
17. No current malaria infection (per rapid diagnostic test)
18. No current Hepatitis B virus infection (per rapid diagnostic test)
19. No current Hepatitis C virus infection (per rapid diagnostic test)
20. No known history or current chronic disease (cancer, kidney disease, or cardiac condition)
21. No known history or current HIV

Among the participants screened, a cross-sectional sample of up to 300 participants (100 per trimester, both anemic and nonanemic) per site will be selected to undergo biomarker-intensive testing. Any method of population-representative sampling may be used within each trimester stratum, provided that selection occurs over the same time period to eliminate time bias.

### Follow-up procedures and data collection

Data will be collected using a harmonized set of forms developed for the PRISMA study [33]. Beyond the routine PRISMA study visit procedures, ReMAPP participants will provide biological specimens to undergo additional laboratory testing (**Table 2**). For hemoglobin, the primary measure of interest, participants will be serially assessed at less than 20 weeks, at 20 weeks, 28 weeks, and 36 weeks gestation and at six weeks postpartum by complete blood count using a venous blood sample and a five-part differential automated hematology analyzer. Additional testing will be done at enrollment, as a part of the screening laboratory tests for the clinically ‘healthy’ subcohort. Biomarker-intensive testing will be done for the sample of 300 participants (100 per trimester), either during their enrollment visit (gestational age less than 14 weeks), 20- week, or 32-week ANC visit. Methods and timing of laboratory assessments will be standardized across sites to ensure comparability and quality.

**Table 2.** Laboratory tests for the ReMAPP study.

| Condition | Assessment | Method |
| --- | --- | --- |

| **Serial hemoglobin tests at <20, 20, 28, and 36 weeks gestation and 6 weeks postpartum** |  |  |
| --- | --- | --- |
| Anemia | Hemoglobin* | Complete blood count using five-part differential hematology analyzer |
| **Screening tests at enrollment (<20 weeks gestation) to identify clinically healthy subcohort** |  |  |
| Iron deficiency | Ferritin | ELISA<sup>1</sup> (Quansys Q-plex™ Human Micronutrient v2 (7-plex)) |
|  | Serum transferrin receptor | ELISA<sup>1</sup> (Quansys Q-plex™ Human Micronutrient v2 (7-plex)) |
| Inflammation | C-reactive protein | ELISA<sup>1</sup> (Quansys Q-plex™ Human Micronutrient v2 (7-plex)) |
|  | Alpha 1-acid glycoprotein | ELISA<sup>1</sup> (Quansys Q-plex™ Human Micronutrient v2 (7-plex)) |
| Communicable diseases | HIV | Rapid diagnostic test |
|  | Malaria | Rapid diagnostic test |
|  | Hepatitis B and C | Rapid diagnostic test |
| Red blood cell defects | Sickle cell disease | HPLC<sup>2</sup> |
|  | Beta (β) and alpha (α) thalassemias | HPLC<sup>2</sup> |
|  | Glucose-6-phosphate dehydrogenase | SD Biosensor Standard G6PD Analyzer |
| **Biomarker-intensive tests at <14, 20, or 32 weeks gestation for random sample** |  |  |
| Iron deficiency | Hepcidin | ELISA<sup>1</sup> |
|  | Total iron-binding capacity | Colorimetric chemistry analyzers |
| Other micronutrient deficiency | Vitamin A serum retinol | HPLC<sup>2</sup> |
|  | Vitamin B12 total cobalamin | ECLIA<sup>3</sup> |
|  | Vitamin B12 holotranscobalamin II | Abbexa HoloTC ELISA<sup>1</sup> |
|  | Folate | Microbiologic assay |
|  | Zinc | ICP-MS<sup>4</sup> or AAS<sup>5</sup> |
| Red blood cell parameters | Red blood cell count | Five-part differential hematology analyzer |
|  | Hematocrit |  |
|  | Mean corpuscular volume |  |
|  | Mean corpuscular hemoglobin concentration |  |
|  | Red cell distribution width |  |
|  | Red blood cell morphological abnormalities | Leishman, Wright-Giemsa, or Giemsa stain for microscopy |
|  | White blood cell morphological abnormalities |  |
|  | Platelet morphological abnormalities |  |
|  | Presence of blood parasites |  |
| Communicable diseases | Syphilis | Rapid diagnostic test |
|  | HIV | Rapid diagnostic test |
|  | Hepatitis B and C | Rapid diagnostic test |
|  | Malaria | Rapid diagnostic test |
|  |  | Placental tissue histopathology |
|  |  | Leishman, Giemsa stain, or Quantitative Buffy Coat for microscopy |
|  | Tuberculosis | GeneXpert (4-module, 8-module, or 24-module) |
|  | Schistosomiasis | Kato-Katz coproscopy, PCR<sup>6</sup> test, or urine filtration |
|  | Soil-transmitted helminths | Kato-Katz coproscopy |
| Non-communicable diseases | Urinalysis | Dipstick |
|  | Renal function | Colorimetric chemistry analyzer |
|  | Liver function | Colorimetric chemistry analyzer |
|  | Blood lead | ICP-MS<sup>4</sup> or AAS<sup>5</sup> |

<sup>1</sup>Enzyme-linked immunoassay (ELISA) <sup>2</sup>High-performance liquid chromatography (HPLC) <sup>3</sup>Electrochemiluminescence (ECLIA) <sup>4</sup>Inductively coupled plasma mass spectrometry (ICP-MS) <sup>5</sup>Atomic Absorption Spectrophotometry (AAS) <sup>6</sup>Polymerase chain reaction (PCR)

### Quality assurance for laboratory tests

Each site laboratory will adhere to the Laboratory Quality Assurance and Quality Control Standard Operating Procedures manual developed specifically for this study. For full blood count measurement, all participating labs will be enrolled onto the United Kingdom National External Quality Assurance Scheme for Hematology for monthly quality assurance [36]. For full blood count and all other analytes, the College of American Pathologists’ External Quality Assessment Program is required [37].

### Sample size estimates

Each site will enroll 1,650 to 2,000 participants in the ReMAPP study, to achieve an overall study sample of 12,000 participants across the six sites. The sample size was selected assuming that about 30% (n=600) will meet the eligibility criteria for the ‘healthy’ subcohort. This sample size will also capture sufficient cases of the primary maternal and newborn outcomes of interest, based on previous regional prevalence estimates, in order to conduct a pooled decision limits analysis with at least 90% power to estimate association between anemia and each outcome.

Our power speculation is based on the distribution of hemoglobin values in pregnancy by gestational age presented in the INTERGROWTH-21^st^ study [18]. For the ‘healthy’ analytical subcohort, a sample size of 600 participants per site allows for single-site estimation of hemoglobin thresholds for any maternal anemia (i.e., 5th percentile) and severe maternal anemia (i.e., 2.5th percentile). As illustrated in **Figure 2**, there will be sufficient precision to distinguish between these two percentiles without overlapping 95% confidence intervals. If sites are unable to identify 600 participants for the healthy cohort, a minimum sample size of 300 participants will be sufficient for single-site hemoglobin threshold estimation.

**Figure 2.**
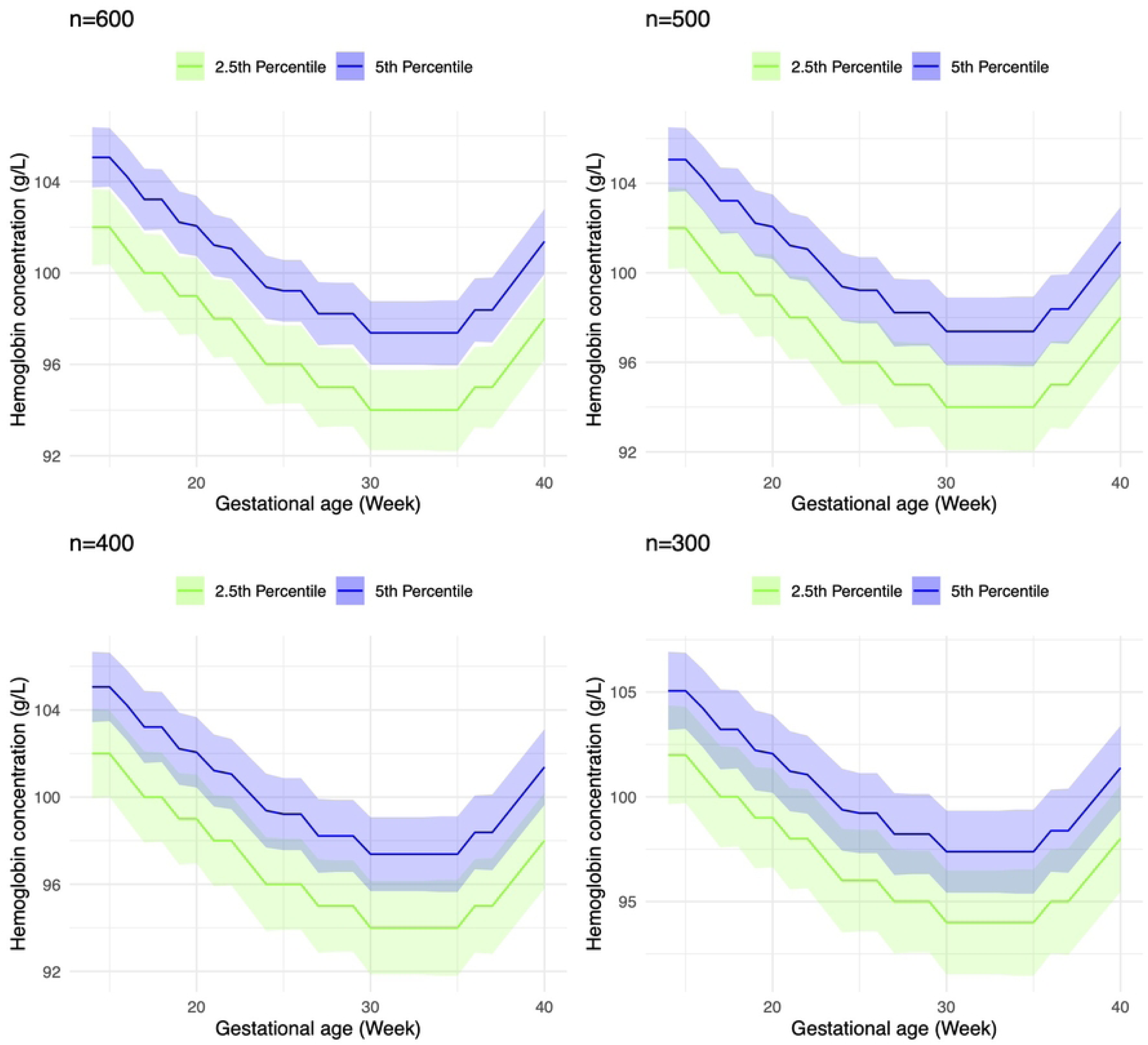
Visualization of sample size estimates with thresholds for any (i.e. 5th percentile) and severe maternal anemia (i.e. 2.5th percentile) at n = 300, 400, 500, 600.

For the cross-sectional sample, n=300 was determined to minimize the burden of additional specimen collection while allowing for exploratory pooled analyses of anemia etiology. The calculation is based on the following assumptions: one third of the participants have a certain contributing factor (e.g., the prevalence of vitamin A insufficiency); 30% of participants without the factor have anemia; and 50% with the factor have anemia. At the two-sided significance level of 95% and 80% statistical power, we can detect a prevalence ratio of 1.7 with a sample size of between 240 and 270 [38]. We inflated the sample to 300 to account for the fact that some contributing factors are less prevalent in certain study sites.

### Participant and public involvement

Participants will be collaboratively involved at multiple phases of the study. During initial protocol workshops, investigators carefully evaluated the additional burden of study participation; for this reason, the size of the biomarker-intensive sample was kept to a minimum and invasive blood draws limited to select visit timepoints. As a part of the translation and validation process for questionnaires to measure fatigue, patients’ experiences were centered and informed scale validation procedures. Participants’ health priorities and experiences will also be explored in corresponding qualitative research about maternal morbidity. Upon completion of the study, we intend to share the main findings with participants and community members via appropriate local dissemination methods. Finally, we will ensure that participants’ contributions to this research are acknowledged in any subsequent reports, presentations, and publications.

### Ethical and safety considerations

This protocol, the informed consent documents and any subsequent modifications will be reviewed and approved by the relevant institutional review board (IRB) and ethics review committee (ERC) responsible for oversight of the study at each site. IRB approvals for this research were received from the following ERCs in each country: Ghana (Ghana Health Service ERC (FWA No. 000200025) and the Kintampo Health Research Center Institutional Ethics Committee, FWA No. 00011103), Kenya (KEMRI Scientific and Ethics Review Unit, 04-10-358- 4166), Zambia (University of Zambia Biomedical Research Ethics Committee, IRB00001131 of IORG0000774 and UNC Biomedical IRB 356795), India (Office of Research, Christian Medical College, Vellore, India Ethics Committee, IRB14553), India (Ethics Review Committee, Society for Applied Studies, New Delhi, SAS/ERC/ReMAPP Study/2022; Department of Health Research, EC/NEW/INST/2022/DL/0140), Pakistan (National Institutes of Health - Health Research Institute, National Bioethics Committee Ref: No.4-87/NBC-962/23/593 and Aga Khan University Ethics Review Committee, 2022-7197-21350), and the United States (Columbia University IRB IRB- AAAU7504; The George Washington University IRB NCR224396; Harvard University IRB IRB- 23-1093). The rights and safety of all study participants will be protected. Written informed consent, assent and parental consent will be sought from all study participants as appropriate prior to enrollment. Site-specific unique identification numbers will be issued to participants instead of names to protect their identity. A limited data set will be stored in a secure cloud server where only trained study staff will have credentials for access. Study participants with abnormal biochemical results will be referred for further clinical assessment and management according to study-specific standard operating procedures.

## ANALYTICAL PLAN

### Aim 1: Establishing hemoglobin clinical decision limits based on adverse outcomes

Aim 1 will establish trimester-specific and postpartum hemoglobin clinical decision limits for any anemia based on associations with adverse maternal, fetal, and infant outcomes (**Table 3**). For this analysis, hemoglobin and outcome data points from all participants will be pooled. Nonparametric relationships between risks of adverse events and hemoglobin levels will be plotted using real data [39]. A series of candidate thresholds in hemoglobin values will be considered and disease risks before and after each threshold will be compared. Thresholds with clinically meaningful differences in the disease risks and statistically sound significance levels will be selected. Clinical decision limits for overall adverse events will be reported, as well as disease-specific clinical decision limits.

**Table 3.** Primary and secondary health outcomes for establishing decision limits.

| Outcome | Definition | Timing |
| --- | --- | --- |
| **Primary outcomes** |  |  |
| Severe maternal outcomes | Composite of maternal deaths, late maternal deaths, and near-misses | Pregnancy through 1 year postpartum |
| Maternal death | Death from any cause related to or aggravated by pregnancy or its management (excluding accidental or incidental causes) | Pregnancy through 42 days postpartum |
| Perinatal depression | Likelihood of depression per the Edinburgh Postnatal Depression Scale | At 20 weeks and 32 weeks gestation; at 6 weeks postpartum |
| Postpartum anemia | Low hemoglobin concentration <12 g/dL | 6 weeks and 6 months |
| Preterm birth | Delivery prior <37 weeks of gestation of a live born infant | <37 weeks gestation |
| Small for gestational age<sup>1</sup> | Birthweight of lower than the sex-specific 10th percentile for each week of gestational age, according to the sex-specific INTERGROWTH reference standards | Delivery |
| Stillbirth | Delivery of a fetus showing no signs of life or >350g birth weight, if gestational age is unavailable | ≥20 weeks gestation |
| **Secondary outcomes** |  |  |
| Late maternal death | Death from any cause related to or aggravated by pregnancy or its management (excluding accidental or incidental causes) | From 42 days to 1 year postpartum |
| Preeclampsia | For participants without preexisting hypertension, defined as gestational hypertension AND gestational proteinuria OR development of severe features. For participants with chronic hypertension, defined as gestational proteinuria OR development of severe features | >20 weeks gestation to 42 days postpartum |
| Hemorrhage | Antepartum hemorrhage (APH): bleeding from or into the genital tract during pregnancy. Postpartum hemorrhage (PPH): estimated blood loss of ≥500 mL, clinical diagnosis of PPH, or receipt of a procedure to manage PPH (including blood transfusion). | ≥24 weeks gestation and prior to delivery (APH), within 24 hours after birth (PPH) |
| Preterm premature rupture of membranes | Rupture of membranes before the onset of labor | <37 weeks of gestation |
| Maternal sepsis | Organ dysfunction resulting from infection | Pregnancy, delivery, and up to 42 days postpartum |
| Maternal fatigue<sup>2</sup> | Decreased capacity for physical and mental activity after childbirth, a persistent lack of energy, impairments in concentration and attention not easily relieved by rest or sleep | At 20 weeks and 32 weeks gestation; at 6 weeks postpartum |
| Low birthweight | Liveborn infant weighing less than 2500 g at birth | Within 72 hours of delivery |
| Neonatal hyperbilirubinemia | Presence of excess bilirubin. Calculated in line with the age-specific total bilirubin thresholds per the American Academy of Pediatrics BiliTool™ algorithm [40] | Birth to 7 days |
| Possible severe bacterial infection | Presence of any of the following signs or symptoms: Not able to feed at all or not feeding well; convulsions; severe chest indrawing; high body temperature ($\geq 38^{\circ}\text{C}$ for <2 months, $\geq 37.5^{\circ}\text{C}$ for 2+ months); low body temperature (less than $35.5^{\circ}\text{C}$); no movement/only when stimulated; fast breathing ($\geq 60$ BPM for <2 months, $\geq 50$ BPM for 2+ months) | Birth to 59 days |
| Neonatal sepsis | Inflammatory response and organ dysfunction following presence of a severe infection from as suspected (by a clinician) or proven (with culture) | Birth to 28 days |
| Infant neurodevelopment<sup>3</sup> | Infant's cognitive, motor, linguistic, and social-emotional development | At 12 months |

<sup>1</sup>Small for gestational age defined as lower than the sex-specific 10th percentile for each week of gestational age per INTERGROWTH standards [41]
<sup>2</sup>Maternal fatigue assessed using the FACIT-Fatigue Scale at 20 weeks and 32 weeks gestation and six weeks postpartum.
<sup>3</sup>Infant neurodevelopment assessed using the Global Scale for Early Development Short Form.

### Aim 2: Establishing hemoglobin reference limits among a healthy population

Aim 2 will establish gestational-age-specific hemoglobin reference limits for any, mild, moderate, or severe anemia in a reference population (i.e., clinically ‘healthy’ subcohort) during pregnancy and within 42 days postpartum [42,43]. This analysis will include hemoglobin data points from all participants eligible for the clinically ‘healthy’ subcohort. Fractional polynomial regression (FPR) will be used to model and visualize the hemoglobin changing curves of different percentiles against gestational age [18]. Sensitivity analysis of the percentile curves predicted by FPR models will be performed excluding one site at a time. If the hemoglobin distributions are similar across sites and between-site between is less than within-site variance, we will pool the data; if not, analyses will remain stratified by site.

Hemoglobin levels below the 5th or 2.5th percentiles at the corresponding time points during pregnancy are typically considered abnormally low and likely indicative of anemia. Therefore, the reference limits are defined as tail percentiles of hemoglobin levels in the reference population. Tail percentiles, such as 5th or 2.5th percentiles, will be modeled as a smooth polynomial function of gestational weeks using hemoglobin levels. With the final validated model, we will estimate the 2.5th, 5th, 95th, and 97.5th percentiles predicted from the FPR curves at specific gestational timepoints from 14 to 40 weeks and postpartum.

### Aim 3: Describing the etiology of anemia in pregnancy

Aim 3 will describe the underlying contributing factors of anemia in pregnancy among a sample of up to 1,800 participants (up to 300 per site), with approximately equal numbers in each trimester. Analyses will be performed on pooled, regional, and site-specific data to identify shared and unique risk factors for anemia. We will quantify the association between contributing factors and anemia (defined using the WHO cutoffs and study-established reference and decision limit thresholds) by calculating relative risks from a generalized linear log-binomial model with robust standard errors. If these models fail to converge, we will use modified Poisson estimates, which produce valid but less efficient estimates of the log-binomial model. We will run separate models for proximal, medial, and distal risk factors (**Figure 3**). Distal risk factors will be included as potential confounders to proximal and medial models. We will estimate the population attributable risk fraction using prevalence of the exposure/risk factor and the relative risk of anemia in the exposed versus unexposed group. Partial population attributable risk will be calculated by fitting multivariate models with log link for anemia adjusting for multiple risk factors. For contributing factors significantly associated with risks of anemia, we will further report the change in continuous hemoglobin concentrations associated with the factor.

**Figure 3.**
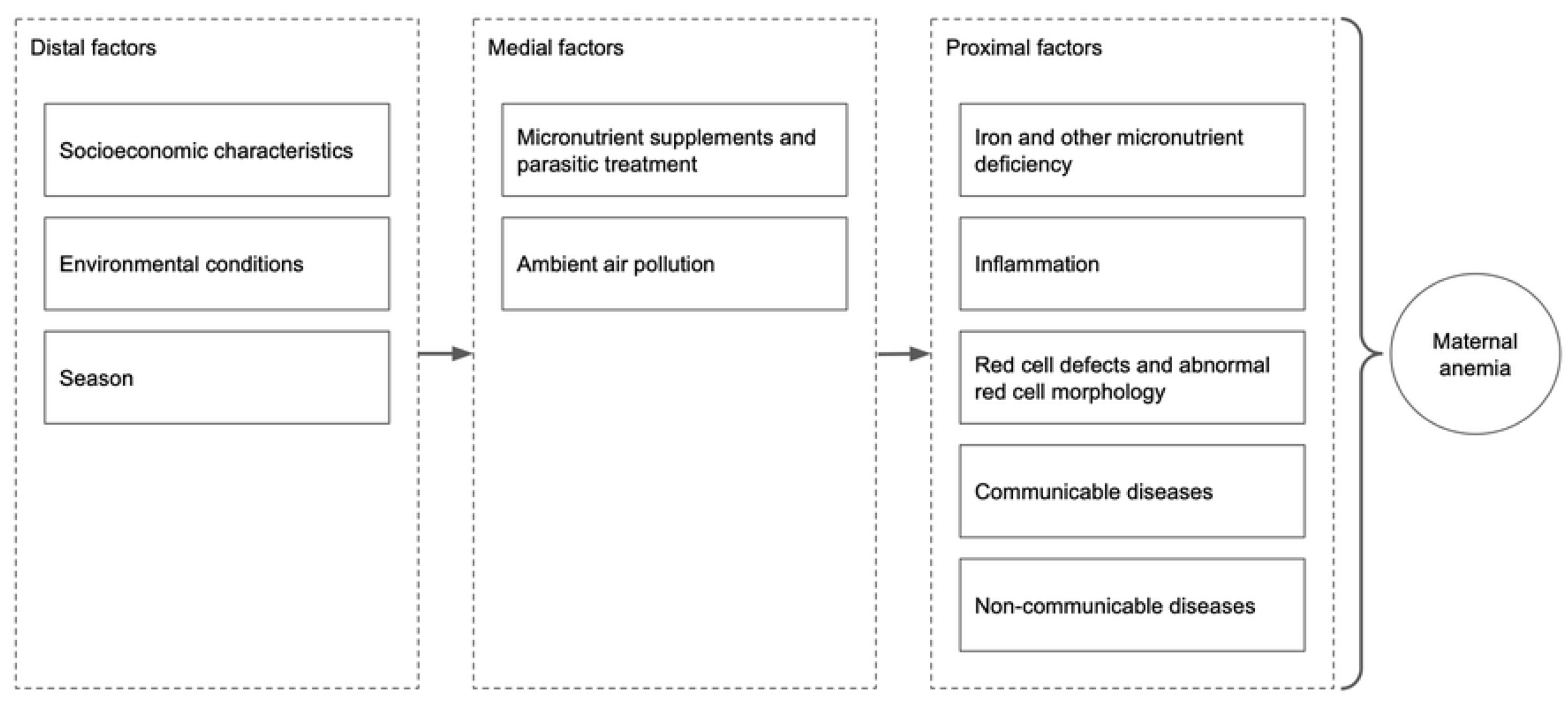
Risk factors of maternal anemia.

### Dissemination plan

The data generated in the course of the study will be reviewed on a regular basis for quality per our established Data Quality Assurance and Quality Control Standard Operating Procedures monthly. Following the end of the study, meetings will be organized at each study site to share results with community members, local authorities, and study participants. Findings will also be shared with local and international stakeholders, including the Bill & Melinda Gates Foundation, WHO, CDC, United States Agency for International Development, and Ministries of Health in each participating country. Abstracts will be developed for dissemination through both local and international scientific conferences and publication in peer-reviewed journals.

## Data Availability

No datasets were generated or analysed during the current study. All relevant data from this study will be made available upon study completion.

## AUTHORS’ CONTRIBUTIONS

Conceptualization: VA, KPA, AGC, ZH, FJ, MPK, SM, VRM, CM, IN, QP, ERS, MBS, and CRS. Funding acquisition: KPA, CT, SN, VA, SJB, IN, MPK, MBS, ERS, CM, and SM. Design of methodology: KPA, CT, ABAK, ERS, QP, LGU, CRS, and BJW. Project administration: VA, DAG, SGB, BF, ASJ, AK, FMM, HM, KO, RSR, WKM, GO, HO, NS, BV, SV, ZAW, JW, and NY. Provision of study materials: VA, DAG, LC, AD, AH, HM, KO, RR, RV, and BV. Supervision: VA, KPA, SJB, ZH, FJ, ASJ, MPK, SM, VRM, CM, WKM, SN, IN, QP, NS, ERS, MBS, CRS, CT, and LGU. Visualization: SGB, QP, and ERS. Writing—original draft preparation: VA, KPA, SGB, QP, and ERS. Writing—review and editing: VA, VA, KPA, SGB, SJB, LC, AGC, AD, DAG, AH, BF, ZH, ASJ, FJ, ABAK, AK, MPK, CM, VRM, HM, SM, IM, FMM, WKM, SN, GO, KO, HO, QP, RSR, NS, ERS, MBS, CRS, CT, LGU, BV, RV, SV, BJW, JW, NY.

## ACKNOWLEDGEMENTS

This study would not be possible without the support of the Bill & Melinda Gates Foundation, specifically from Laura Lamberti and Sun-Eun Lee. The study design greatly benefited from the expertise of the ReMAPP Technical Advisory Group members, who were generous with their time and expertise: Lisa Rogers, Maria Elena Jefferds, Denish Moorthy, Melissa Fox Young, Nicholas Kassebaum, Eric Ohuma, and Bosede Afolabi. ReMAPP investigators are grateful to the extensive network of individuals and organizations who contributed to the design of this study, including but not limited to biostatisticians, laboratory technicians, obstetricians, community health workers, field data collectors, and research partners.

## REFERENCES

1. Kassebaum NJ, Jasrasaria R, Naghavi M, Wulf SK, Johns N, Lozano R, et al. A systematic analysis of global anemia burden from 1990 to 2010. Blood. 2014;123: 615–624.

2. Anemia in Pregnancy: ACOG Practice Bulletin, Number 233. Obstet Gynecol. 2021;138: e55–e64.

3. Stevens GA, Paciorek CJ, Flores-Urrutia MC, Borghi E, Namaste S, Wirth JP, et al. National, regional, and global estimates of anaemia by severity in women and children for 2000-19: a pooled analysis of population-representative data. Lancet Glob Health. 2022;10: e627–e639.

4. Safiri S, Kolahi A-A, Noori M, Nejadghaderi SA, Karamzad N, Bragazzi NL, et al. Burden of anemia and its underlying causes in 204 countries and territories, 1990-2019: results from the Global Burden of Disease Study 2019. J Hematol Oncol. 2021;14: 185.

5. Yang Y, Hou Y, Wang H, Gao X, Wang X, Li J, et al. Maternal Thyroid Dysfunction and Gestational Anemia Risk: Meta-Analysis and New Data. Front Endocrinol. 2020;11: 201.

6. Deng Y, Steenland K, Sinharoy SS, Peel JL, Ye W, Pillarisetti A, et al. Association of household air pollution exposure and anemia among pregnant women: Analysis of baseline data from “Household Air Pollution Intervention Network (HAPIN)” trial. Environ Int. 2024;190: 108815.

7. Yadav G, Chambial S, Agrawal N, Gothwal M, Kathuria P, Singh P, et al. Blood lead levels in antenatal women and its association with iron deficiency anemia and adverse pregnancy outcomes. J Family Med Prim Care. 2020;9: 3106–3111.

8. Ortiz-Garcia NY, Cipriano Ramírez AI, Juarez K, Brand Galindo J, Briceño G, Calderon Martinez E. Maternal Exposure to Arsenic and Its Impact on Maternal and Fetal Health: A Review. Cureus. 2023;15: e49177.

9. Azami M, Badfar G, Khalighi Z, Qasemi P, Shohani M, Soleymani A, et al. The association between anemia and postpartum depression: A systematic review and meta-analysis. Caspian J Intern Med. 2019;10: 115–124.

10. Kang SY, Kim H-B, Sunwoo S. Association between anemia and maternal depression: A systematic review and meta-analysis. J Psychiatr Res. 2020;122: 88–96.

11. Young MF, Oaks BM, Tandon S, Martorell R, Dewey KG, Wendt AS. Maternal hemoglobin concentrations across pregnancy and maternal and child health: a systematic review and meta-analysis. Ann N Y Acad Sci. 2019;1450: 47–68.

12. Sukrat B, Wilasrusmee C, Siribumrungwong B, McEvoy M, Okascharoen C, Attia J, et al. Hemoglobin concentration and pregnancy outcomes: a systematic review and meta-analysis. Biomed Res Int. 2013;2013: 769057.

13. Wedderburn CJ, Ringshaw JE, Donald KA, Joshi SH, Subramoney S, Fouche J-P, et al. Association of Maternal and Child Anemia With Brain Structure in Early Life in South Africa. JAMA Netw Open. 2022;5: e2244772.

14. Steer PJ. Maternal hemoglobin concentration and birth weight. Am J Clin Nutr. 2000;71: 1285S–7S.

15. Yip R. Significance of an abnormally low or high hemoglobin concentration during pregnancy: special consideration of iron nutrition. Am J Clin Nutr. 2000;72: 272S–279S.

16. Amburgey OA, Ing E, Badger GJ, Bernstein IM. Maternal hemoglobin concentration and its association with birth weight in newborns of mothers with preeclampsia. J Matern Fetal Neonatal Med. 2009;22: 740–744.

17. Young MF, Oaks BM, Rogers HP, Tandon S, Martorell R, Dewey KG, et al. Maternal low and high hemoglobin concentrations and associations with adverse maternal and infant health outcomes: an updated global systematic review and meta-analysis. BMC Pregnancy Childbirth. 2023;23: 264.

18. Ohuma EO, Young MF, Martorell R, Ismail LC, Peña-Rosas JP, Purwar M, et al. International values for haemoglobin distributions in healthy pregnant women. EClinicalMedicine. 2020;29-30: 100660.

19. Chang S-C, O’Brien KO, Nathanson MS, Mancini J, Witter FR. Hemoglobin concentrations influence birth outcomes in pregnant African-American adolescents. J Nutr. 2003;133: 2348–2355.

20. Yip R. Significance of an abnormally low or high hemoglobin concentration during pregnancy: special consideration of iron nutrition. Am J Clin Nutr. 2000;72: 272S–279S.

21. Garcia-Casal MN, Pasricha S-R, Sharma AJ, Peña-Rosas JP. Use and interpretation of hemoglobin concentrations for assessing anemia status in individuals and populations: results from a WHO technical meeting. Ann N Y Acad Sci. 2019;1450: 5–14.

22. Guideline on haemoglobin cutoffs to define anaemia in individuals and populations. World Health Organization; 2024 Mar. Available: https://www.who.int/publications/i/item/9789240088542

23. Chaparro CM, Suchdev PS. Anemia epidemiology, pathophysiology, and etiology in low- and middle-income countries. Ann N Y Acad Sci. 2019;1450: 15–31.

24. World Health Organization. Iron Deficiency Anaemia: Report of a Study Group. World Health Organization Technical Report Series; 1959. Report No.: 182. Available: https://apps.who.int/iris/bitstream/handle/10665/40447/WHO_TRS_182.pdf

25. World Health Organization. WHO Scientific Group on Nutritional Anaemias & World Health Organization. Nutritional anaemias : report of a WHO scientific group [meeting held in Geneva from 13 to 17 March 1967]. 1968. Available: https://apps.who.int/iris/handle/10665/40707

26. Natvig K. Studies on hemoglobin values in Norway. V. Hemoglobin concentration and hematocrit in men aged 15-21 years. Acta Med Scand. 1966;180: 613–620.

27. Kilpatrick GS HRM. The prevalence of anaemia in the community. British Medical Journal. 1961. pp. 778–782.

28. De Leeuw KM Lowenstein L Hsieh YS. Iron deficiency and hydremia in normal pregnancy. Medicine. The Wiliams & Wilkins Co.; 1966. pp. 291–315.

29. Sturgeon P. Studies of iron requirements in infants. III. Influence of supplemental iron during normal pregnancy on mother and infant. A. The mother. British Journal of Haematology. 1959.

30. CDC Criteria for Anemia in Children/Childbearing-Aged Women. 9 Jun 1989 [cited 9 Mar 2021]. Available: https://wonder.cdc.gov/wonder/prevguid/p0000169/p0000169.asp

31. World Health Organization. WHO recommendations on antenatal care for a positive pregnancy experience. 2016. Available: https://apps.who.int/iris/bitstream/handle/10665/250796/9789241549912-eng.pdf;jsessionid=B18B1C73CA88F51F6DD4326F3906382B?sequence=1

32. World Health Organization. Haemoglobin concentrations for the diagnosis of anaemia and assessment of severity. Vitamin and Mineral Nutrition Information System; 2011. Available: https://www.who.int/vmnis/indicators/haemoglobin.pdf

33. Smith ER, Baumann SG, Mores C, Tawiah C, Asante KP, Mazumder S, et al. PRISMA Maternal & Newborn Health Study. In: Open Science Foundation [Internet]. OSF; 1 Oct 2024 [cited 2 Oct 2024]. Available: https://osf.io/qckyt/

34. World Health Organization. Nutritional Anaemias: Tools for Effective Prevention and Control. 2017.

35. Suchdev PS, Namaste SML, Aaron GJ, Raiten DJ, Brown KH, Flores-Ayala R, et al. Overview of the Biomarkers Reflecting Inflammation and Nutritional Determinants of Anemia (BRINDA) Project. Adv Nutr. 2016;7: 349–356.

36. Full Blood Count (FBC) Programme. In: UK NEQAS [Internet]. 6 Mar 2017 [cited 15 Jul 2024]. Available: https://ukneqas.org.uk/programmes/result/?programme=full-blood-count-%28fbc%29

37. Proficiency Testing (PT)/External Quality Assessment (EQA) Programs Process. In: College of American Pathologists [Internet]. 26 Jan 2018 [cited 15 Jul 2024]. Available: https://www.cap.org/laboratory-improvement/laboratories-outside-the-usa/proficiency-testing-pt-external-quality-assessment-eqa-programs/proficiency-testing-pt-external-quality-assessment-eqa-programs-process

38. Kelsey JL. Methods in Observational Epidemiology. Monographs in Epidemiology and; 1996.

39. Lai Y. On the adaptive partition approach to the detection of multiple change-points. PLoS One. 2011;6: e19754.

40. Kemper AR, Newman TB, Slaughter JL, Maisels MJ, Watchko JF, Downs SM, et al. Clinical Practice Guideline Revision: Management of Hyperbilirubinemia in the Newborn Infant 35 or More Weeks of Gestation. Pediatrics. 2022;150. doi:10.1542/peds.2022-058859

41. Fernandes M, Villar J, Stein A, Staines Urias E, Garza C, Victora CG, et al. INTERGROWTH-21st Project international INTER-NDA standards for child development at 2 years of age: an international prospective population-based study. BMJ Open. 2020;10: e035258.

42. Sikaris K. Application of the stockholm hierarchy to defining the quality of reference intervals and clinical decision limits. Clin Biochem Rev. 2012;33: 141–148.

43. Ozarda Y, Sikaris K, Streichert T, Macri J, IFCC Committee on Reference intervals and Decision Limits (C-RIDL). Distinguishing reference intervals and clinical decision limits - A review by the IFCC Committee on Reference Intervals and Decision Limits. Crit Rev Clin Lab Sci. 2018;55: 420–431.

